# A Multi-Cohort Study of Immunoglobulin G Glycans in Newly Diagnosed Inflammatory Bowel Disease Patients Reveals Accelerated Biological Aging

**DOI:** 10.64898/2026.04.10.26349930

**Authors:** Konstantinos Flevaris, Irena Trbojević-Akmačić, David Goh, Juproop Singh Lalli, Frano Vučković, Marija Ćapin Vilaj, Jerko Štambuk, Jasminka Krištić, Anika Mijakovac, Nick Ventham, Rahul Kalla, Anna Latiano, Natalia Manetti, Dalin Li, Dermot P.B. McGovern, Nicholas A. Kennedy, Vito Annese, Gordan Lauc, Jack Satsangi, Cleo Kontoravdi

## Abstract

**Background and Aims:** Alterations in immunoglobulin G (IgG) N-glycosylation are implicated in inflammatory bowel disease (IBD); however, the robustness of IgG glycan signatures across IBD cohorts with diverse demographics and geographic origins remains underexplored. We aimed to determine whether compositional data analysis (CoDA) and machine learning (ML) can identify IBD-related IgG N-glycan signatures and whether these signatures capture disease-associated acceleration of biological aging.

**Methods:** We analyzed the IgG glycome profiles of 1,367 plasma samples collected from healthy controls (HC), symptomatic controls (SC), and people with newly diagnosed Crohn’s (CD), and ulcerative colitis (UC) across four cohorts (UK, Italy, United States, and Netherlands). IgG glycosylation was analyzed by ultra-high-performance liquid chromatography, yielding 24 total-area-normalized glycan peaks (GPs). Analyses were performed using cross-sectional data obtained at baseline. CoDA-powered association analyses were used to identify disease-related effects on GPs while controlling for demographic covariates. ML models were trained and evaluated to assess generalizability to unseen cohorts and demographic subgroups, with a focus on discrimination and reliability.

**Results:** Across all cohorts, people with IBD demonstrated accelerated biological aging as quantified by the GlycanAge index. This was accompanied by consistent reductions in IgG galactosylation, with effects partially modulated by age. Classification models trained on glycomics and demographics achieved robust discrimination (AUROC≈0.80) between non-IBD (HC+SC) and IBD across cohorts.

**Conclusion:** These findings reveal accelerated biological aging in people with IBD and support the translational potential of IgG glycans as biomarkers and a novel route toward clinically interpretable personalized risk estimates.

## Introduction

Inflammatory bowel disease (IBD), comprising Crohn’s disease (CD) and ulcerative colitis (UC), affects approximately 1 in 200 individuals in developed countries^1^, imposing an annual healthcare burden estimated at over €5 billion across Europe^2^. The current diagnostic gold standard, namely colonoscopy with ileoscopy and histological examination of multiple biopsies^3^, is invasive and unsuitable for frequent monitoring. Accordingly, there is an ongoing need for robust, non-invasive biomarkers that capture disease activity and progression. Existing serum and fecal markers provide a partial solution to this; C-reactive protein (CRP) reflects systemic inflammation but shows variable sensitivity across IBD subtypes^4^ while fecal calprotectin (FCP) reliably detects mucosal inflammation but lacks disease specificity^5^. Although both can aid in monitoring disease activity and treatment response, and while elevated FCP during clinical remission associates with an increased risk of relapse, their overall ability to predict individual long-term disease course and complications remains to be established^6,7^. These limitations have motivated the discovery of novel, non-invasive molecular biomarkers that can integrate systemic inflammatory and gastrointestinal immune signals^8,9^ towards precision medicine approaches in IBD.

Glycobiology holds promise for advancing biomarker research in various disease conditions by revealing disease predisposition, progression, and potentially identifying molecular therapeutic targets^10,11^. N-glycans are structurally diverse polysaccharides that are co- and post-translationally attached to proteins with well-documented immunological significance and direct implication in disease pathophysiology^12–14^. Importantly, the N-glycan profile of a given protein can vary with disease state^15,16^. Immunoglobulin G (IgG) glycosylation is of particular interest, as alterations in the N-glycan structures attached to the IgG crystallizable fragment (Fc) region modulate its effector functions, including antibody-dependent cellular cytotoxicity (ADCC) and complement-dependent cytotoxicity (CDC)^17^. A large body of work has demonstrated that IgG N-glycosylation patterns are altered in various inflammatory and autoimmune diseases^18,19^, supported by genome-wide association studies (GWAS), revealing pleiotropic genetic effects between IgG glycosylation traits and disease susceptibility^20^.

Several studies have reported IBD-associated changes in IgG glycosylation, most notably a decrease in IgG galactosylation (increased agalactosylated IgG species) in patients with active disease^21–25^. This shift to agalactosylated IgG results in a more pro-inflammatory phenotype. Such glycosylation changes could thus not only contribute to IBD pathogenesis but also serve as robust biomarkers of immune-mediated inflammation. By profiling IgG N-glycans, one can capture an integrated snapshot of the systemic immune milieu that complements existing blood and fecal markers. In fact, emerging evidence suggests that IgG glycosylation signatures may precede clinical disease onset^25^, highlighting their potential value in early diagnosis and risk prediction. Nevertheless, to support the translation of these findings into clinically actionable biomarkers, it is essential to establish generalizable IgG N-glycan signatures that can reliably distinguish IBD patients from healthy individuals and those with non-inflammatory conditions across diverse populations. This is particularly challenging due to the complex interplay of genetic and environmental factors influencing glycosylation, the technical expertise required for high-throughput glycan analysis^26,27^, and the statistical challenges in analyzing the resulting data^28,29^.

In this work, we analyzed serum IgG N-glycan profiles from four geographically distinct cohorts of newly diagnosed IBD using high-throughput ultra-high-performance liquid chromatography (UHPLC), and applied compositional data analysis (CoDA)^29,30^ and machine learning (ML) methods^31^ to identify robust disease-associated glycan signatures. We show that the IgG N-glycan alterations consistently distinguishing non-IBD from IBD are characterized by reduced galactosylation in an age-dependent manner, and are associated with accelerated glycan-derived biological age as quantified by the GlycanAge index^32^. Furthermore, prediction models trained on pooled discovery cohorts also retained robust discrimination in held-out validation cohorts. Collectively, our analysis highlights the translational potential of IgG N-glycan profiling as a biomarker platform for inflammatory disease and a route toward clinically interpretable personalized IBD risk estimates.

## Materials and Methods

### Clinical Samples and Ethical Considerations

The dataset used in this study was generated within the European Commission-funded IBD-BIOM research program, which commenced on October 1, 2012 and concluded on September 30, 2016. The primary objective of the IBD-BIOM consortium was to identify and validate biomarkers for IBD by integrating clinical data with high-throughput molecular technologies, including glycomics^23,33–40^. Samples were collected from four geographically distinct prospective cohorts, namely the Edinburgh cohort (n=527) from the United Kingdom (*UK*), the Cedars-Sinai cohort (n=472) from the United States (*US*), the Florence cohort (n=400) from Italy (*IT*), and the Maastricht cohort (n=183) from the Netherlands (*NL*). All cohorts were collected with the approval of the local ethics committees and informed consent was obtained from all participants. Phenotype was defined using the Montreal classification at the last follow-up^34^. The study included newly diagnosed CD or UC patients without previous administration of therapy, as well as healthy controls (HC) and symptomatic controls (SC) as non-inflammatory comparators. Key demographic and clinical characteristics of the study population are summarized in Table 1.

**Table 1.**
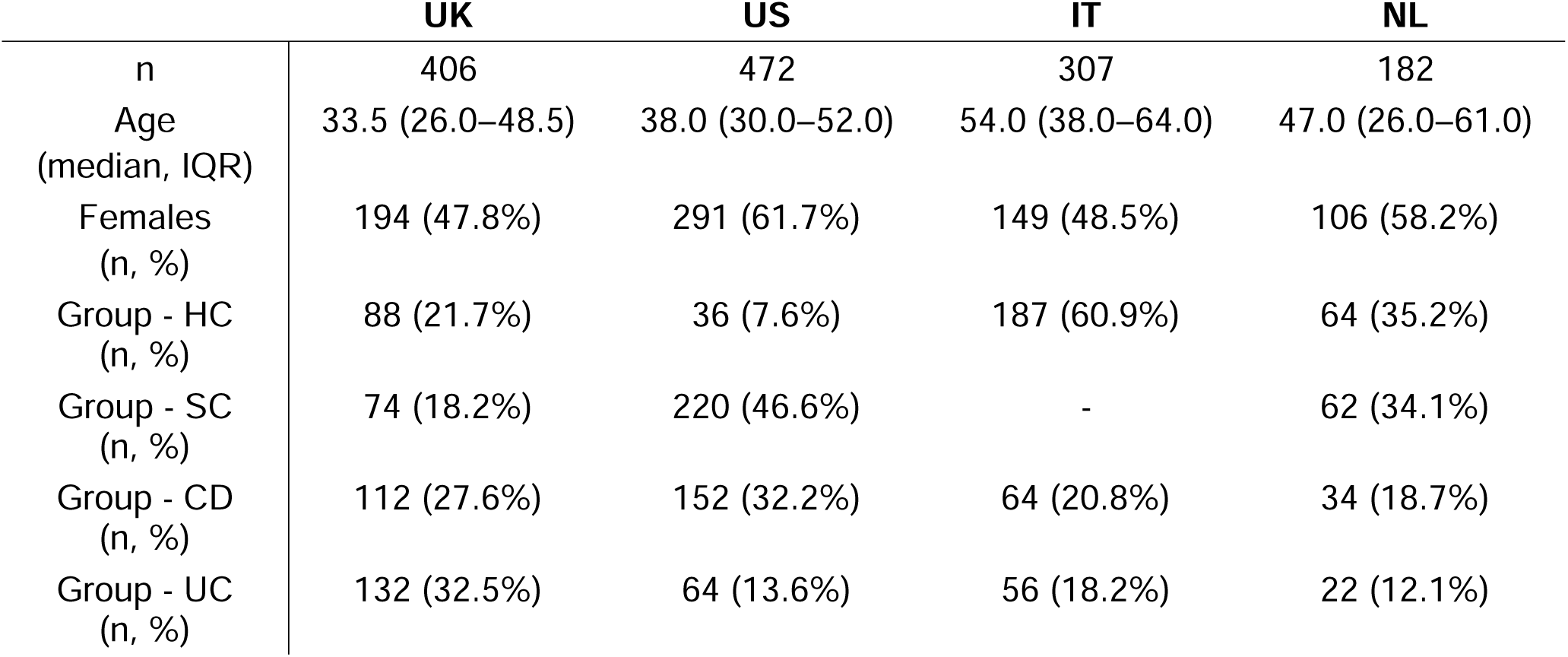
Demographic and clinical characteristics of the study cohorts.

|  | UK | US | IT | NL |
| --- | --- | --- | --- | --- |
| n | 406 | 472 | 307 | 182 |
| Age<br>(median, IQR) | 33.5 (26.0–48.5) | 38.0 (30.0–52.0) | 54.0 (38.0–64.0) | 47.0 (26.0–61.0) |
| Females<br>(n, %) | 194 (47.8%) | 291 (61.7%) | 149 (48.5%) | 106 (58.2%) |
| Group - HC<br>(n, %) | 88 (21.7%) | 36 (7.6%) | 187 (60.9%) | 64 (35.2%) |
| Group - SC<br>(n, %) | 74 (18.2%) | 220 (46.6%) | - | 62 (34.1%) |
| Group - CD<br>(n, %) | 112 (27.6%) | 152 (32.2%) | 64 (20.8%) | 34 (18.7%) |
| Group - UC<br>(n, %) | 132 (32.5%) | 64 (13.6%) | 56 (18.2%) | 22 (12.1%) |

### Sample Preparation and Data Preprocessing

Each cohort included plasma-derived batch-corrected total-area-normalized relative abundances of 24 total IgG GPs, measured by high-throughput UHPLC, as previously described^44^. The GPs correspond to distinct N-glycan structures or groups of structures, which are summarized in Table S1 using their respective Oxford notation^41^ and GlyTouCan identifier^42^. The GPs were total-area-normalized within each sample, such that the sum of relative abundance values equalled 100%. As such, the GPs were treated as compositional data, meaning non-negative vectors that represent relative proportions of parts of a whole, thus encoding relative information^29,30^. All analyses were performed using cross-sectional data obtained at baseline. Unless otherwise stated, GPs were preprocessed using the scale-uncertain centered log-ratio (CLR) transformation with a default uncertainty parameter (γ) of 0.1^29^ prior to analysis. All CoDA techniques throughout this study were performed using Python (v3.12) using the *glycowork*^43^ package.

### Statistical Analysis

The similarity between glycan profiles across all disease groups was visualized by applying principal component analysis (PCA) to beta-diversity matrices. To account for potential confounding factors, the effects of age and sex were first removed through residualization^44^. Specifically, CLR-transformed GPs were regressed on age and sex covariates using Ridge regression, providing residualized values that isolate variation unrelated to these demographic variables by removing pertinent linear signal. Beta-diversity was quantified by calculating pairwise Aitchison distances (i.e., Euclidean distances after CLR transformation) between samples, following the approach of Bennet et al.^29^. Classical multidimensional scaling (CMDS) was subsequently applied to the distance matrices to derive coordinates suitable for PCA^45^. Principal components (PCs) were extracted, and the first two dimensions were examined to assess the separation between disease groups. Analyses were conducted separately for each cohort to account for cohort-specific variation. To complement visual assessments, quantitative clustering metrics, namely the Dunn index and mean silhouette width, were calculated^29^.

Biological age was quantified using the GlycanAge index, which was calculated according to Krištić et al.^32^ (see Supplementary Material for more information). Biological age acceleration (BAA) was defined as the difference between GlycanAge and the chronological age of each individual at the time of sampling.

Association analysis was carried out to estimate disease-related effects on CLR-transformed GPs. For each GP, we fitted an ordinary least squares (OLS) model with disease status, age (centered at 40, with an additional quadratic term), and sex as predictors, including disease-by-age and age-by-sex interactions and cohort indicators. Heteroscedasticity-consistent standard errors were used throughout. As a sensitivity analysis, each model was re-fitted separately in females and males, and corresponding disease coefficients were compared between strata using a Wald-type z-test on the difference in estimates.

P-values were adjusted for multiple testing using a two-stage Benjamini-Hochberg procedure to control the false discovery rate (FDR)^29^. In addition, permutation-based FDR (1,000 permutations) was computed for the disease main-effect comparisons to provide a non-parametric confirmation of significance. Unless otherwise specified, results with FDR-adjusted p-values less than 0.05 were considered statistically significant. All statistical analyses throughout this study were performed using Python (v3.12) using the *glycowork*^43^, *statsmodels*^46^, and *scipy*^47^ packages.

### Machine Learning Analysis

Predictive modeling was performed to classify patients with IBD from those with SC and from HC using GPs, age, and sex as inputs. A suite of ML pipelines was developed, each combining a specific feature representation with a classification algorithm. Feature representations included raw GPs, CLR-transformed GPs, and glycomotifs (GlyCmp-transformed GPs) generated using GlyCompareCT^48^. For classification, logistic regression (LR) and XGBoost^49^ (XB) were employed.

Model generalization was evaluated using a nested leave-one-cohort-out (LOCO) cross-validation (CV) framework, in which models were trained on three cohorts and tested on the remaining held-out cohort. The three cohorts in the inner LOCO-CV loop were further split into training and test sets using stratified sampling to preserve the distribution of disease status, age, sex, and cohort membership. The inner LOCO-CV test sets serve as an independent in-sample evaluation of model performance with respect to known populations, while the held-out cohort provides an out-of-sample performance assessment with respect to new populations. To reduce the risk of learning cohort-specific or demographically imbalanced patterns, two mechanisms were incorporated. First, to promote equal representation of demographic groups during training, sample weights inversely proportional to their prevalence in the training data were applied across eight disease-age-sex strata (age dichotomized at 40 years). Second, all ML pipelines were fit with explicit regularization to limit model complexity and prevent the learning of spurious correlations that may reduce transportability (see Table S3). Subgroup analyses were then conducted to evaluate performance across the age-sex strata in both in-sample and out-of-sample settings. The entire LOCO-CV procedure was repeated five times using different random seeds to ensure numerical stability and robustness of the estimated performance. Model performance was quantified using AUROC and log loss, with the latter serving as a proper scoring rule that jointly reflects discrimination and the reliability of predicted probabilities^50^. Clinically relevant metrics, such as sensitivity and specificity, are also reported. All ML analyses were performed using Python (v3.12) with the scikit-learn^51^ package.

## Results

### IBD Patients Possess Distinct Global IgG N-glycome Profiles Compared to Non-Inflammatory Conditions

There was a clear distinction between IgG glycan profiles in IBD and non-IBD individuals. The distinction between both CD and UC, and also HC and SC, was less pronounced. To investigate between-group similarity and overall structure in the data, we examined global patterns in IgG N-glycome variation using PCA on beta-diversity matrices across all cohorts. Figure S1 and Figure S2 present the cohort-specific biplots under two labeling schemes: (i) a binary representation combining HC and SC into a single “Non-IBD” group vs. IBD (CD and UC) and (ii) a multiclass representation distinguishing all four disease groups (HC, SC, CD, UC). In both instances, the first two principal components (PCs) explained a substantial portion of the variance in IgG N-glycome profiles (approximately 65%), with the binary representation yielding clearer separation between Non-IBD and IBD groups compared to the multiclass one across all cohorts, as reflected in the clustering metrics (Table S2). The Dunn index remained relatively stable across cohorts, ranging from 0.074 to 0.126 under multiclass labeling and from 0.074 to 0.128 under binary labeling, indicating comparable worst-case separation in both settings. The mean silhouette width, while modest, shifted from marginally negative values (−0.042 to −0.023) in the multiclass setting to positive values (0.046 to 0.078) under binary labeling, reflecting enhanced within-cluster cohesion when contrasting individuals with and without IBD. Together, these results indicate that dichotomising by systemic inflammation yields clearer separation and reduces unnecessary variance.

### IBD Patients Exhibit Age-Dependent Changes in IgG Galactosylation in Core Fucosylated Structures

Disease-associated effects on CLR-transformed IgG glycan peaks at the reference age of 40 years are summarized in Figure 1 and Table 2. Compared to HC, SC showed only two FDR-significant differences, namely in GP4/FA2 and GP9/FA2[3]G1, indicating overall similarity of the SC and HC circulating IgG glycomes. In contrast, both CD and UC exhibited a significant compositional shift characterized by enrichment in agalactosylated structures. These effects were broadly directionally concordant between CD and UC and generally larger in CD, supporting the presence of a shared glycomic response to systemic inflammation and underscoring the difficulty of distinguishing the two IBD phenotypes based on glycan profiles alone. At the level of individual features, digalactosylated glycans exhibited the most prominent reductions in IBD, encompassing both high-abundance species such as FA2G2 (GP14) and related G2-containing structures, as well as several low-abundance glycans showing appreciable relative shifts on the scale-uncertain CLR scale (Table 2). In parallel, agalactosylated glycans displayed consistent increases across both CD and UC, most notably FA1 (GP1) and FA2 (GP4). Changes in sialylated glycans were more heterogeneous. In CD, several sialylated structures were reduced relative to HC, including FA2G2S1 (GP18), A2BG2S2 (GP22), FA2G2S2 (GP23), and FA2BG2S2 (GP24), whereas UC showed a mixed pattern. When considered in the context of relative abundance, changes in highly abundant glycans, including FA2 (GP4) and members of the G1/G2 and G2/S1 groups (e.g., FA2[6]G1/GP8, FA2G2/GP14, FA2G2S1/GP18), appear to drive the overall disease-associated compositional signature, whereas large effects observed in low-abundance glycans contribute less to the global glycan distribution.

**Figure 1.**
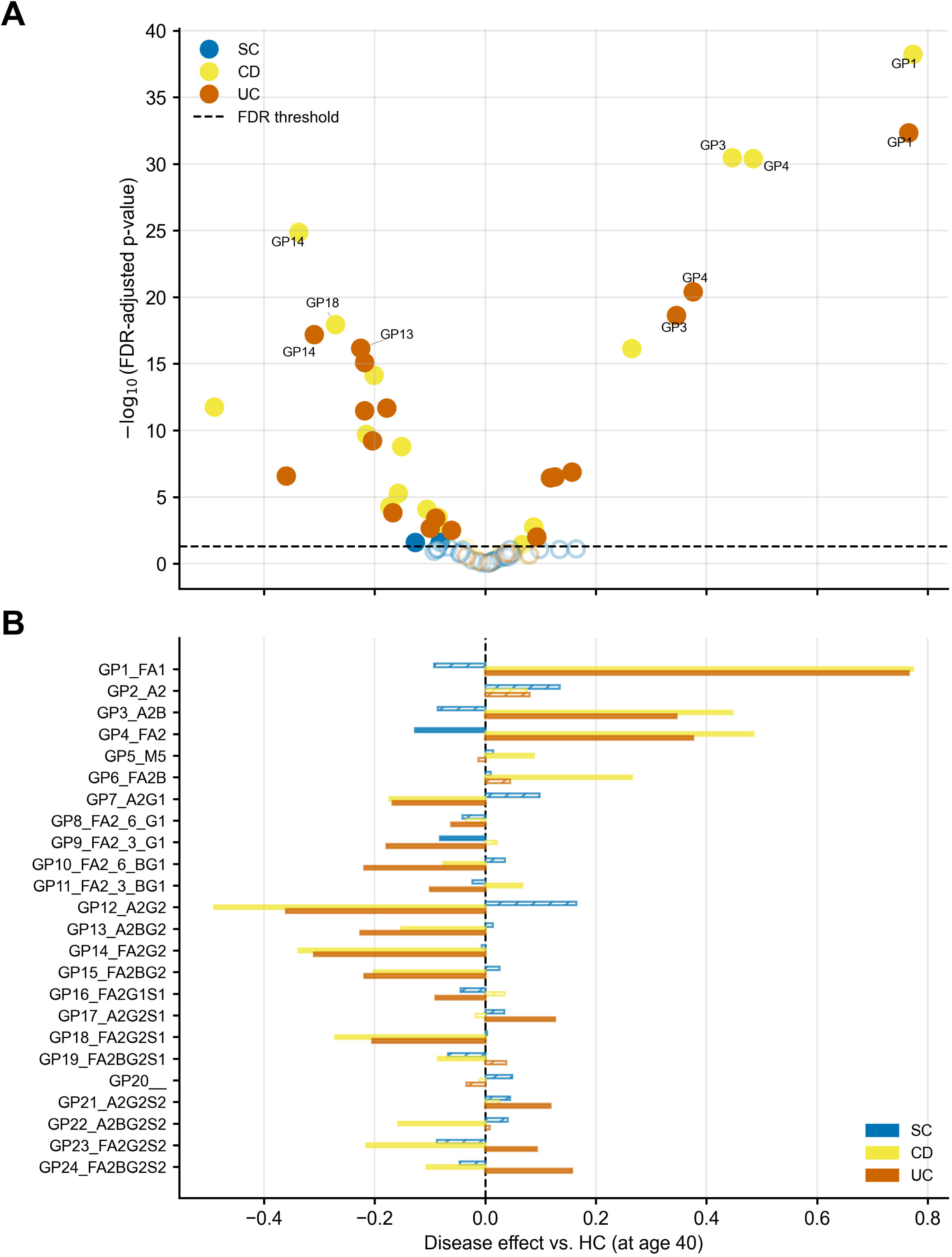
Disease-associated alterations (main effects) in IgG N-glycans in SC and IBD patients. (A) Volcano plot of fixed-effect coefficients from linear models comparing SC, CD and UC to HC for each GP. The dashed horizontal line marks the significance threshold (FDR-adjusted p < 0.05, Benjamini–Hochberg correction). (B) Horizontal bar plot showing the estimated effect sizes for each GP in SC, CD, and UC compared to HC. Effect sizes represent CLR-transformed log-ratio shifts relative to the geometric mean of all glycans, adjusted for age, sex, and cohort, evaluated at age 40. Positive values indicate higher relative glycan abundance compared to HC; negative values indicate lower abundance. Solid bars denote significant effects (FDR-adjusted p < 0.05); hatched bars denote non-significant effects.

**Table 2.** Key IgG N-glycan changes associated with IBD phenotypes. Shown are regression coefficients (β) and 95% confidence intervals from compositional CLR-transformed GPs comparing Crohn’s disease (CD) and ulcerative colitis (UC) to healthy controls (HC). Positive coefficients indicate relative enrichment and negative coefficients relative reduction of the corresponding glycan.

| Glycan (GP) | CD $\beta$ (95% CI) | UC $\beta$ (95% CI) |
| --- | --- | --- |
| **FA1 (GP1)** | +0.77 [0.66, 0.89] | +0.77 [0.64, 0.89] |
| **A2B (GP3)** | +0.45 [0.37, 0.52] | +0.35 [0.27, 0.42] |
| **FA2 (GP4)** | +0.48 [0.40, 0.57] | +0.38 [0.30, 0.45] |
| **A2G2 (GP12)** | -0.49 [-0.63, -0.35] | -0.36 [-0.50, -0.22] |
| **FA2G2 (GP14)** | -0.34 [-0.40, -0.27] | -0.31 [-0.38, -0.24] |
| **FA2BG2 (GP15)** | -0.20 [-0.25, -0.15] | -0.22 [-0.27, -0.17] |
| **FA2G2S1 (GP18)** | -0.27 [-0.33, -0.21] | -0.20 [-0.27, -0.14] |

Age-based interaction analyses were directionally aligned with the main disease effects, but this pattern was substantially more pronounced in CD than in UC (Figure S3). In CD, 11 of 24 glycans showed significant disease-by-age interactions, indicating a modest attenuation of the disease-associated agalactosylated signature with increasing age. Specifically, agalactosylated glycans such as FA1 (GP1) showed negative age interactions (β=−0.016 yr⁻¹, 95% CI [−0.024, −0.009]), whereas several galactosylated structures showed positive age interactions, most notably A2G2 (GP12; β=+0.020 yr⁻¹, 95% CI [0.012, 0.029]), together with smaller positive shifts in related G1/G2-containing glycans. In UC, age interactions were fewer and weaker, with only three glycans reaching significance, all showing negative coefficients for agalactosylated structures, including FA1 (GP1; β =−0.012 yr⁻¹, 95% CI [−0.019, −0.005]), A2B (GP3; β=−0.007 yr⁻¹, 95% CI [−0.013, −0.002]), and FA2 (GP4; β =−0.007 yr⁻¹, 95% CI [−0.013, −0.0002]). No significant age interactions were observed in SC. Overall, these findings indicate that age modestly modulates the IBD-associated glycan signature, with a clearer shift towards higher galactosylation in older CD patients. In all cases, the interaction magnitudes remained small relative to the corresponding main disease effects. In sex-stratified sensitivity analyses, no male-female difference in the estimated disease coefficients remained significant after FDR correction, suggesting that the main disease associations were broadly consistent across sexes.

### The IgG N-glycome Profiles of IBD Patients Across Cohorts Are Reflective of Accelerated Biological Aging

The IgG N-glycome profiles of IBD patients exhibited accelerated biological aging compared with those of individuals without IBD (Figure 2). Across all cohorts, the IBD group shows consistently higher BAA than non-IBD individuals (median BAA: UK 13.5 vs. 5.9; US 11.9 vs. 2.8; IT 12.8 vs. −1.1; NL 10.0 vs. 0.0), with highly significant comparisons in every cohort (all p-adj < 0.01). The largest increase was seen in CD (median BAA: UK 18.2; US 13.0; IT 19.5; NL 9.7), and the difference in BAA values between HC and CD was significant in all cohorts. BAA differences between HC and UC were also significant across cohorts, whereas differences between HC and SC individuals were less discernible and cohort-dependent (significant only in UK, p-adj = 0.038). The latter aligned with previous observations about the similarity of IgG N-glycomes between HC and SC (i.e., non-IBD). Notably, the IT cohort did not include any SC individuals, which resulted in negative BAA values. This indicates that the GlycanAge of the HC samples, and thus of the non-IBD samples, was lower than their respective chronological age. Consequently, this suggests that, on average, HC individuals in the IT cohort were “healthier” than HC individuals in the other cohorts included in this study.

**Figure 2.**
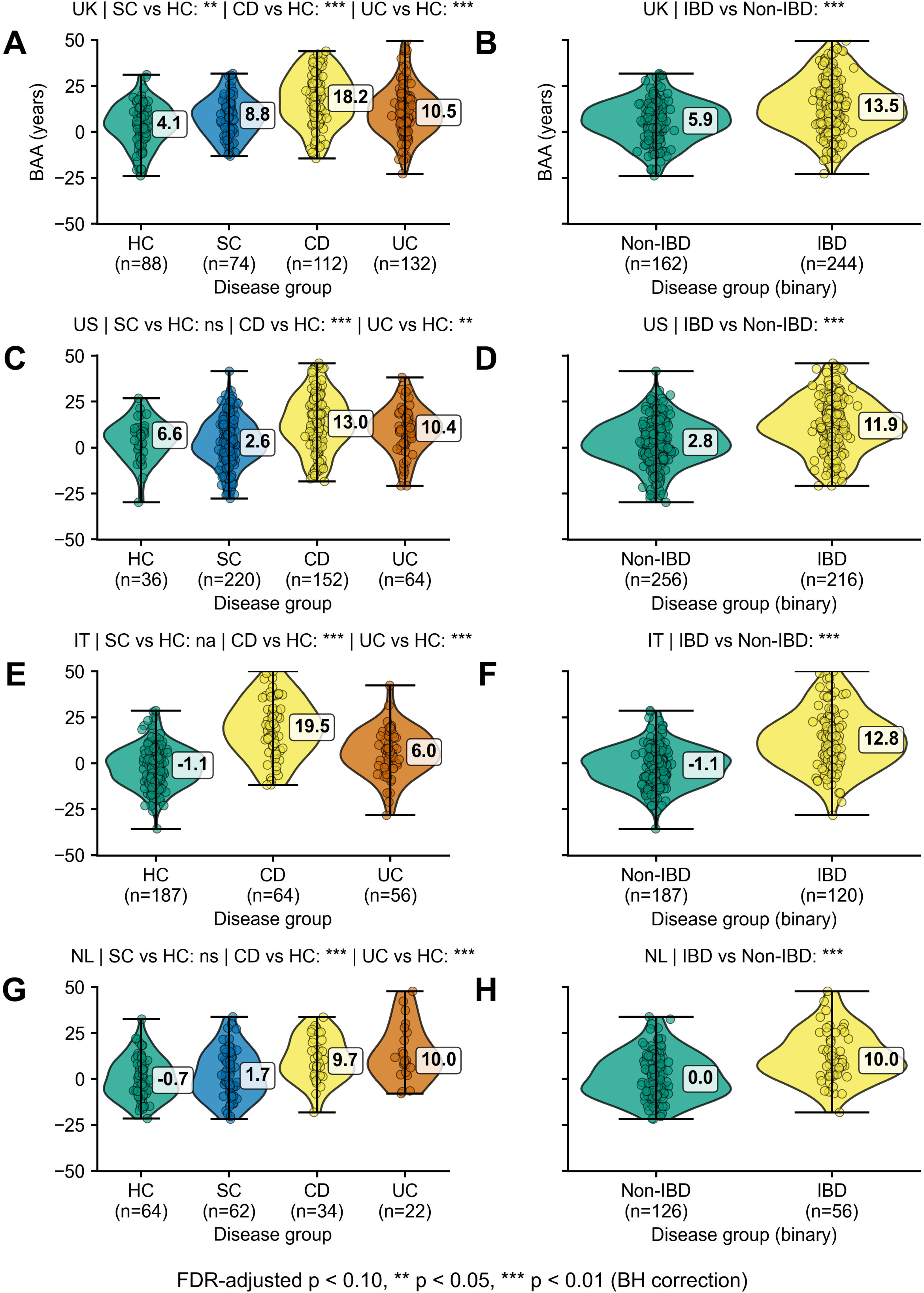
Differences in biological age acceleration across cohorts and disease groups. Panels are organized by cohort in row pairs, namely (A)-(B) for UK, (C)-(D) for US, (E)-(F) for IT, and (G)-(H) for NL. Within each cohort, the left panel shows all disease groups (i.e., HC, SC, CD, UC) and the right panel shows the binary comparison Non-IBD (HC+SC) vs. IBD (CD+UC). All pairwise comparisons use two-sided Mann-Whitney U tests with Benjamini-Hochberg FDR adjustment within each cohort. Significance is indicated by star symbols corresponding to FDR-adjusted p-value thresholds of 0.10, 0.05, and 0.01. Non-significant comparisons are denoted by *ns* and non-applicable comparisons are denoted by *na*.

### IgG N-glycan Signatures Robustly Discriminate IBD from Non-IBD Across Cohorts

We evaluated the suitability of IgG N-glycan profiles to discriminate IBD from Non-IBD (HC & SC) using various ML pipelines under a nested LOCO CV framework. This design was used to quantify absolute classification performance and to assess generalization to unseen populations at two levels: (i) overall performance across cohorts, and (ii) subgroup-specific performance within demographic groups (i.e., predefined age-sex strata).

Across all pipelines, generalization performance was stable, with small to moderate gaps between in-sample and out-of-sample evaluations depending on the held-out cohort (Figure 3). LR- and XB-based models exhibited consistent behavior across UK, US, IT, and NL cohorts despite differing inductive biases, indicating that the predictive glycomic signal is largely model-agnostic. Likewise, raw, CLR-transformed, and GlyCmp features performed comparably overall, although pointwise generalization gaps revealed cohort-specific trends. UK and NL consistently showed minimal or slightly favorable external performance for both AUROC and log loss, indicating that these cohorts are closely aligned with the distributions learned during training. In contrast, IT displayed the largest degradations across nearly all pipelines, while US showed smaller but directionally inconsistent shifts. The former likely reflects the absence of SC samples, which necessitates merging HC and SC during training and may yield decision boundaries that do not generalize optimally at test time. At the same time, there could be cohort-specific glycomics and clinical differences that are not captured by the feature space and could be driven by unobserved clinical factors. GlyCmp representations showed slightly larger variability relative to raw and CLR features, particularly in IT, although the magnitude of these effects remained modest. Despite these cohort-specific differences, generalization gaps converged around zero when aggregated across pipelines, supporting the conclusion that the biological signal discriminating IBD from Non-IBD is robust to modeling choices and largely stable across the cohorts in this study.

**Figure 3.**
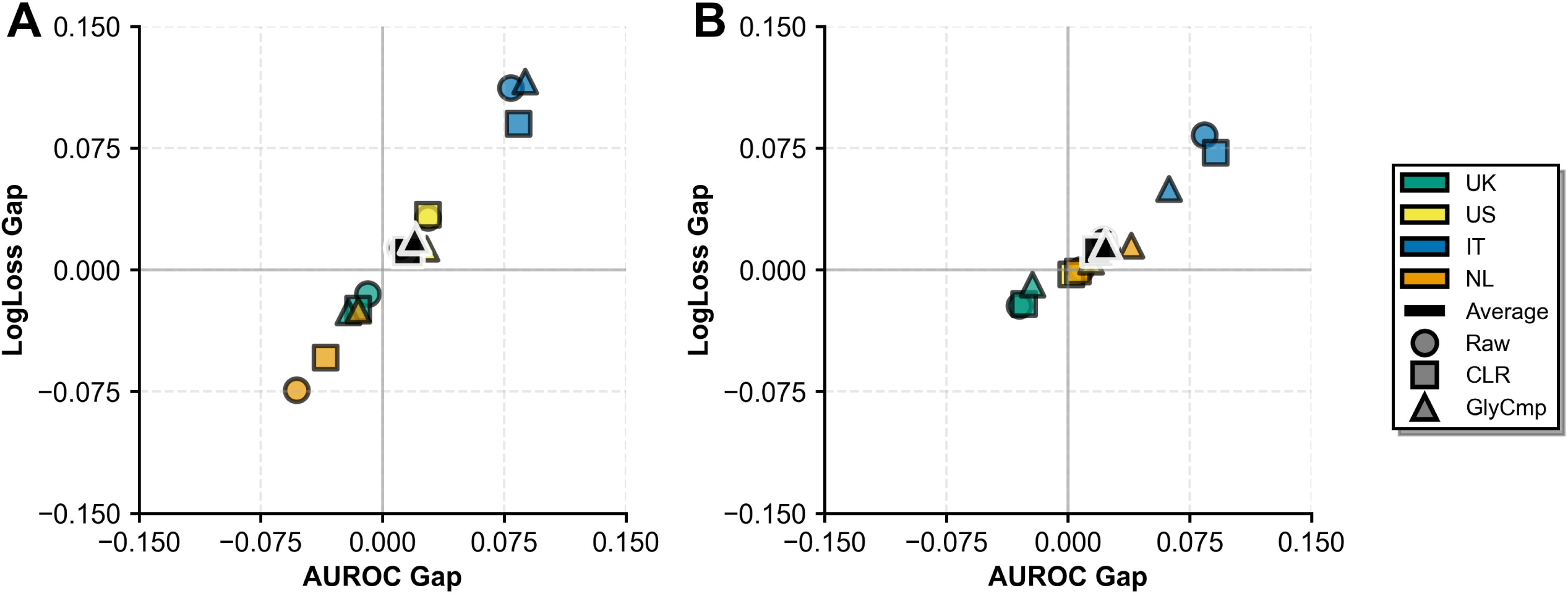
Overall generalization gaps for ML prediction models classifying Non-IBD vs. IBD under LOCO-CV. The generalization gaps in AUROC and log loss between in-sample and out-of-sample evaluations are shown for all held-out cohorts and feature representation methods in (A) LR and (B) XB models. Each point represents one ML pipeline evaluated on one held-out cohort, with colors indicating the held-out cohort and shapes indicating the feature representation method (raw GPs, CLR-transformed GPs, or GlyCmp-transformed GPs). The intersection of the horizontal and vertical lines indicates perfect generalization (i.e., zero gap) for both metrics.

Absolute out-of-sample performance (Figure 4; Table S4) further supports these conclusions and highlights clinically relevant trade-offs. Averaged across held-out cohorts, LR pipelines achieved higher sensitivity (≈0.64-0.68) than XB (≈0.59-0.61), while both approaches showed comparable specificity (≈0.75-0.80). When considered alongside AUROC and log loss, LR models delivered performance that was comparable to, or better than, XB while maintaining more balanced error profiles, indicating greater robustness to cohort-level heterogeneity. CLR features yielded modest improvements over raw inputs for both model classes, whereas GlyCmp representations yielded lower AUROCs and higher log loss values, indicating a potential loss of discriminative information through motif-level aggregation. Importantly, this likely arises from the intrinsically limited structural diversity of IgG N-glycans (predominantly bi-antennary species) rather than from the GlyCompare methodology^28^ itself. Consistent with earlier analyses, IT was the most challenging cohort across all pipelines. Overall, these results indicate that raw GP features combined with LR provide a strong, low-complexity baseline that is robust to inter-cohort variability.

**Figure 4.**
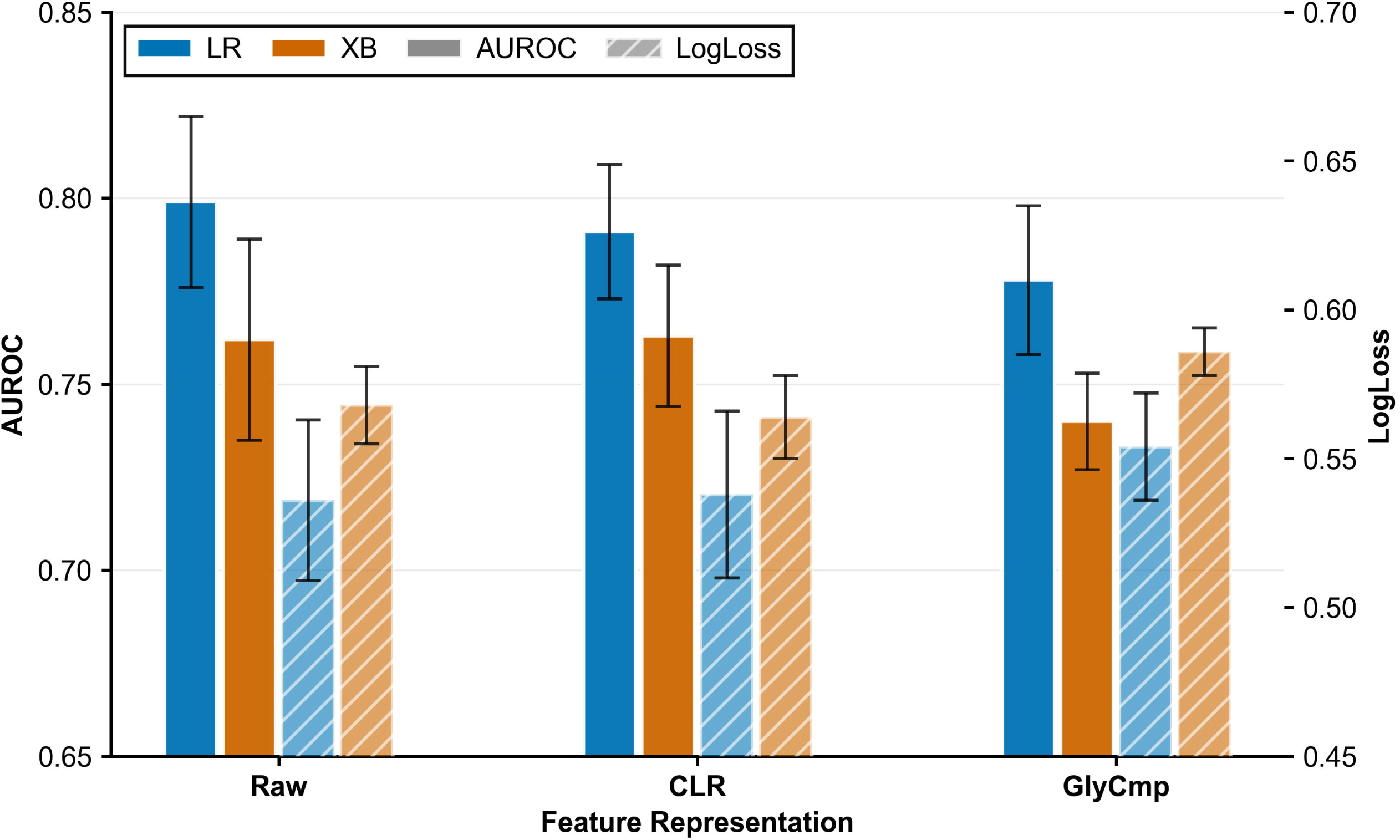
Overall out-of-sample performance for ML prediction models classifying Non-IBD vs. IBD under LOCO-CV. Absolute out-of-sample performance in terms of AUROC and log loss is shown across all held-out cohorts for each feature representation method (raw GPs, CLR-transformed GPs, or GlyCmp-transformed GPs) and model class (LR or XB). Each bar represents the mean out-of-sample performance across held-out cohorts for a given model class and feature representation, and error bars indicate the standard error across held-out cohorts. Colors indicate model class and hatching indicates the evaluation metric.

Across demographic strata, subgroup generalization was broadly stable, with AUROC and log loss gaps mostly clustering near zero for both LR and XB pipelines (Figure 5). No age-sex subgroup showed substantial degradation when moving from in-sample to out-of-sample evaluation. For LR, younger strata exhibited slightly larger variability in AUROC and log loss gaps, particularly for raw and GlyCmp features, whereas CLR-transformed inputs generally pulled subgroup averages closer to the origin, indicating more reliable generalization across held-out cohorts. XB pipelines showed a similar pattern with tighter clustering overall, indicating marginally more stable cross-cohort behavior while preserving the same relative subgroup and feature-ordering trends observed for LR. Overall, these results indicate the absence of systematic demographic bias, with modest subgroup-specific fluctuations attenuated by CLR preprocessing.

**Figure 5.**
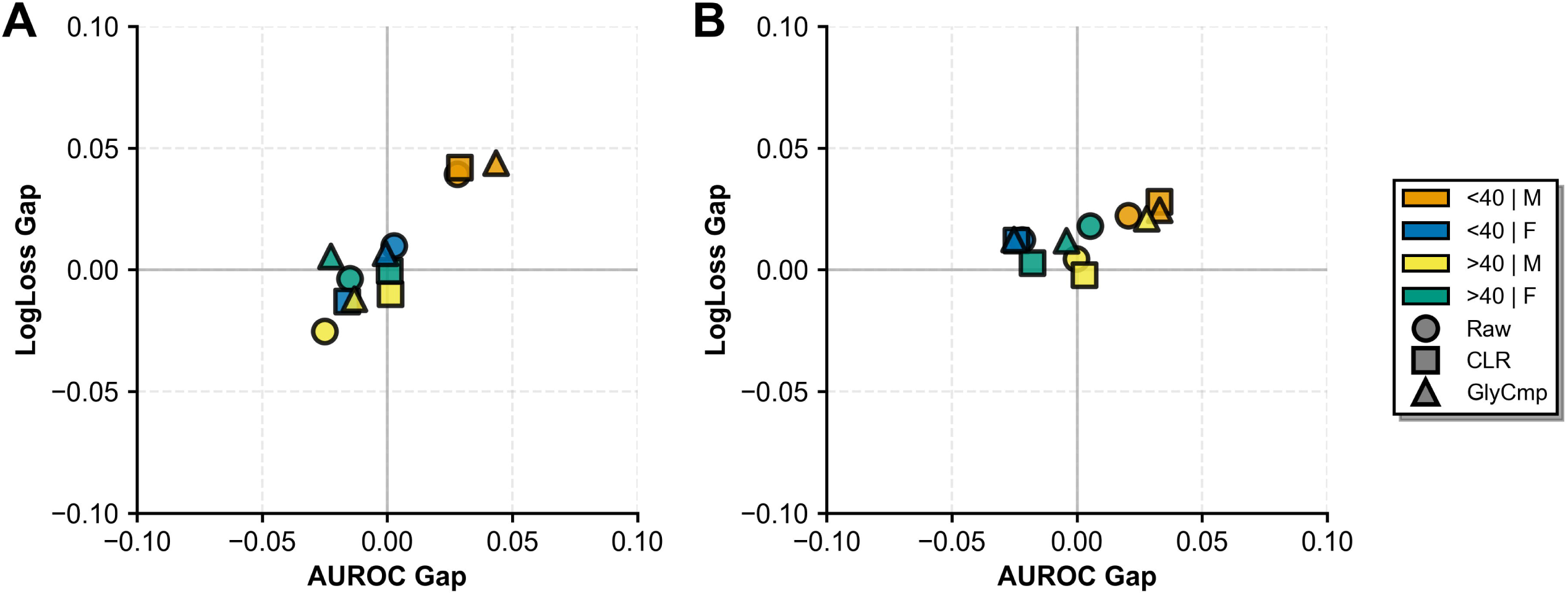
Subgroup generalization gaps for ML prediction models classifying Non-IBD vs. IBD across age-by-sex strata under LOCO-CV. Age was binarized as ≤40 years and >40 years. The generalization gaps in AUROC and log loss between in-sample and out-of-sample evaluations are averaged across all held-out cohorts for each feature representation method in (A) LR and (B) XB models. Each point represents the cohort-averaged generalization gap of an ML pipeline within a specific demographic stratum, with colors indicating the demographic stratum and shapes indicating the feature representation method (raw GPs, CLR-transformed GPs, or GlyCmp-transformed GPs). The intersection of the horizontal and vertical lines indicates perfect generalization (i.e., zero gap) for both metrics.

Absolute out-of-sample performance across demographic subgroups (Figure S4; Tables S5-S8) remained in the moderate-to-high discrimination range and log loss values indicative of stable probabilistic calibration. Younger males and females exhibited the strongest performance, with AUROCs typically exceeding 0.80 for both LR and XB across raw and CLR representations, whereas older subgroups, particularly older females, showed modestly reduced accuracy, most notably with GlyCmp features. CLR preprocessing yielded small but consistent improvements within strata, while GlyCmp features produced lower AUROC and higher log loss across model classes, mirroring overall out-of-sample trends. Importantly, LR and XB showed closely matched performance in all demographic groups, indicating that the underlying glycomic signal generalizes similarly across age and sex strata and supporting the robustness of glycan-based classifiers to demographic variation.

Finally, the entire training and evaluation process was repeated to assess whether CD patients could be reliably distinguished from UC patients using LR based on their glycomics profiles and demographics (Table S9). Notably, while discrimination ability was moderate (AUROC≈0.70-0.73), prediction confidence was not satisfactory (log loss≈0.62-0.66). These findings suggest that, although IgG glycosylation captures some disease-specific signal, it is insufficient to support reliable CD-UC differentiation in this setting.

## Discussion

In this study, a multi-center IgG N-glycomics dataset comprising 1,367 plasma samples was assembled from four geographic cohorts (*UK*, *US*, *IT*, *NL*) under the IBD-BIOM consortium. Four phenotype groups were considered: HC, CD, UC, and SC, the latter likely representing individuals with functional bowel conditions such as irritable bowel syndrome (IBS), but without formal diagnosis. Demographic characteristics differed across disease groups, with variation in age, sex, and cohort membership, underscoring the need to account for covariate effects in downstream analyses. To emulate a discover-then-validate workflow commonly employed in epidemiological research, we implemented a LOCO-CV evaluation scheme, where each cohort served as an unseen test cohort, thereby enabling unbiased evaluation of ML prediction models and enhancing confidence in their generalizability to unseen heterogeneous populations.

Even though changes in IgG glycosylation have been reported multiple times in the past^21–25^, this work should not be viewed as a simple replication study, since in addition to being the largest study of IgG N-glycosylation in IBD to date, contrary to all past studies, it was performed on four geographically distinct cohorts of carefully recruited newly diagnosed, treatment-naive patients, which demonstrated that the observed changes may reflect disease pathophysiology and are not an effect secondary to therapy. We report a decrease in IgG galactosylation, pointing particularly to reductions in the relative abundances of digalactosylated and core fucosylated N-glycan structures (e.g., FA2G2) counteracted by proportional increases in agalactosylated species (e.g., FA2). Our analysis employing CoDA techniques allows the identification of a set of GPs that jointly shift in relative abundance, rather than focusing on isolated glycans. This perspective captures the interconnected nature of glycan biosynthesis and its modulation in disease states. These findings align with prior reports in IBD, as well as across autoimmune diseases, cancer, and inflammatory conditions^52^, suggesting that IgG galactosylation likely serves as a general biomarker/modulator of systemic inflammation, consistent across multiple cohorts. We identified sialylation changes as well; however, their effect sizes were less pronounced and likely a consequence of altered galactosylation, as galactose residues serve as substrates for sialic acid addition in the canonical biosynthetic network. Regarding differences between CD and UC, our results indicate largely concordant glycan alterations in both conditions, with CD exhibiting slightly larger effect sizes. This aligns with existing research in CD and glycomics, where lower IgG-Fc galactosylation has been shown to precede CD onset and even contribute to the initiation of inflammation^25^. This difficulty in distinguishing UC from CD is consistent with observations across multiple omics disciplines, including DNA methylation analyses^40^, which have consistently reported substantial molecular overlap between the two conditions.

A key finding of this work is the consistent identification of accelerated biological aging in patients with IBD at diagnosis, as quantified by the GlycanAge index^32^. Elevated GlycanAge was observed across all cohorts and populations, underscoring that accelerated biological aging is a reproducible feature of IBD rather than a cohort-specific finding. To our knowledge, this is the first report of GlycanAge-based acceleration in biological aging in IBD cohorts spanning multiple geographic locations. GlycanAge has been used as a robust biomarker for monitoring changes in biological age^53^, even under pharmacological interventions^54^. Importantly, these findings are consistent with other biological aging frameworks implicated in IBD. Blood-biomarker-based aging measures, particularly PhenoAge^55^, have shown strong associations with IBD risk^56^. In large population-based analyses from the UK Biobank, PhenoAge acceleration (the difference between PhenoAge and chronological age) was associated with a substantially increased incident risk of IBD, CD, and UC, with each standard deviation increase conferring approximately a one-third to one-half higher disease risk^56^. DNA methylation-based epigenetic clocks, including Horvath and GrimAge^57^, have also linked age acceleration to inflammatory states and immune dysregulation in IBD^40,58,59^. Recent evidence in CD indicates that even among patients in long-term clinical remission on combination biologic and immunomodulator therapy, approximately 15-20% exhibit marked epigenetic age acceleration (>5 years)^60^, implying that accelerated biological aging may persist despite apparent disease control. Within this broader landscape, the GlycanAge-related findings in this study appear particularly striking; whereas epigenetic clocks often identify pronounced age acceleration in subsets of patients, GlycanAge-enabled BAA captured a broadly shared shift at diagnosis, supported by highly consistent data across cohorts. This may reflect the unique sensitivity of IgG glycosylation patterns to immune activation and inflammatory burden, features that are central to IBD pathophysiology. Together, these data support a unifying model in which IBD is characterized by early and measurable acceleration of biological aging, rooted in immune dysregulation and inflammation, detectable across multiple molecular layers, and evident at or before clinical diagnosis.

CoDA techniques were central to our data analysis methodology, addressing the compositional nature of glycomics data and enabling robust statistical inference. While CoDA has only recently been applied in the context of N-glycomics^29^, our study incorporates these approaches to plasma-derived IgG N-glycome data across multiple geographically and demographically distinct cohorts, thereby broadening their applicability to large, heterogeneous clinical datasets. As demonstrated by Bennett et al.^29^, CoDA methods mitigate inflated Type I error rates arising from spurious correlations in glycomics data, an issue that is likely to become increasingly pronounced as larger, multi-cohort studies emerge and data are pooled across diverse populations. It is key to note that the CoDA-powered statistical framework employed here, while enabling the detection of coordinated glycan shifts in disease states, does not directly elucidate the underlying, causal, biological mechanisms driving these alterations. Future studies integrating glycomics with transcriptomics or proteomics could provide deeper insights into the regulatory pathways involved.

In the last decade, notable advancements have been made in building prediction models for disease classification using glycomics data^52^. A wide array of disease conditions has been investigated with sample sizes varying substantially across studies, ranging from modest discovery cohorts to large-scale datasets involving a few thousand individuals from diverse geographical and ethnic populations^61–64^. Nevertheless, and likely due to the relative novelty of glycomics as a high-throughput omics technology, existing studies predominantly focus on single cohorts with limited demographic diversity, which may restrict generalizability. Additionally, the uncertainty in predicted probabilities is not routinely assessed and has only recently been examined in glycomics-based prediction models^57^. Ignoring this uncertainty may lead to the over- or underestimation of disease risk in certain individuals, thereby compromising clinical decisions about which individuals should be prioritized for further testing. Building on these observations, we propose a methodology that aligns with best practices in clinical prediction modeling in gastroenterology^65,66^ through assessment of generalization and robustness of our ML models across diverse demographic subgroups and geographically distinct cohorts.

Given that the use of high-throughput N-glycan data for disease risk prediction is a nascent field, we believe the work herein can support the development of reliable prediction models that are equitable across different populations. Potential improvements include the integration of additional omics modalities (e.g., proteomics, metabolomics) alongside patient-level information, including ethnicity, BMI, and co-morbidities, to enhance predictive performance and robustness. At the same time, incorporating N-glycans from multiple protein sources (e.g., total plasma, acute-phase proteins) could provide complementary information that may improve disease classification. Efforts to enhance the diversity of glycomics datasets by measuring multiple proteins from the same individuals are underway^67^ and total plasma N-glycans have already shown promise as biomarkers in IBD^68^. Taken together, this study supports the translational potential of IgG N-glycan profiling as a biomarker platform for inflammatory disease and a route toward clinically interpretable risk estimates.

## Declaration of Generative AI and AI-assisted technologies in the writing process

During the preparation of this work the authors used ChatGPT (OpenAI) in order to assist with language refinement. After using this tool, the authors reviewed and edited the content as needed and take full responsibility for the content of the publication.

## Data Availability

All analysis code is publicly available at the GitHub repository: https://github.com/kf120/ibd-biom-glycoda. The data that support the findings of this study are available on reasonable request from the corresponding author.

## Acknowledgments

KF thanks the Department of Chemical Engineering at Imperial College London for his scholarship.

## Grant Support

This work was supported by the European Union’s Horizon 2020 Research and Innovation Programme through the SynHealth project (grant no. 101159018), and by the European Union’s Seventh Framework Programme through the IBD-BIOM project (grant no. 305479).

## Disclosures

GL is the founder and owner of Genos Ltd., a private research organization specializing in high-throughput glycomic analysis and holding several patents in this field. IT-A, FV, MĆV, JŠ, JK, AM are employees of Genos Ltd. GL is also the founder and owner of Genos Glycoscience Ltd., a spin-off company of Genos Ltd. that commercializes its scientific discoveries, and a co-founder and Chief Scientific Officer of GlycanAge Ltd., which markets the GlycanAge biological age test. DPBM is a consultant for Mirador Therapeutics and holds patents for anti-TL1A therapy in IBD. JS has received grant funding from the EC, CCUK, and the Helmsley Trust for biomarker studies in IBD at the University of Oxford, and from the EC for IBD-BIOM and IBD-CHARACTER at the University of Edinburgh. NV, RK, AL, NM, DL, NAK, VA, KF, DG, JSL, and CK have nothing to disclose.

## Author Contributions

Konstantinos Flevaris, KF (Conceptualization; Data curation; Formal Analysis; Investigation; Methodology; Software; Visualization; Writing – original draft, Writing – review & editing)

Irena Trbojević-Akmačić, IT-A (Formal Analysis; Investigation; Methodology; Writing – review & editing)

David Goh, DG (Formal Analysis; Investigation; Methodology; Software)

Juproop Singh Lalli, JSL (Formal Analysis; Investigation; Methodology)

Frano Vučković, FV (Formal Analysis; Investigation; Methodology)

Marija Ćapin Vilaj, MĆV (Formal Analysis; Investigation; Methodology)

Jerko Štambuk, JŠ (Formal Analysis; Investigation; Methodology)

Jasminka Krištić, JK (Formal Analysis; Investigation; Methodology)

Anika Mijakovac, AM (Investigation; Writing – review & editing)

Nick Ventham, NV (Investigation; Writing – review & editing)

Rahul Kalla, RK (Investigation; Writing – review & editing)

Anna Latiano, AL (Investigation; Writing – review & editing)

Natalia Manetti, NM (Investigation; Writing – review & editing)

Dalin Li, DL (Investigation; Writing – review & editing)

Dermot P.B. McGovern, DPBM (Investigation; Writing – review & editing)

Nicholas A. Kennedy, NAK (Investigation; Writing – review & editing)

Vito Annese, VA (Conceptualization; Project administration; Resources; Writing – review & editing)

Gordan Lauc, GL (Conceptualization; Investigation; Methodology; Project administration; Resources; Writing – review & editing)

Jack Satsangi, JS (Conceptualization; Investigation; Methodology; Project administration; Resources; Writing – review & editing)

Cleo Kontoravdi, CK (Conceptualization; Investigation; Methodology; Project administration; Resources; Writing – review & editing)

## Data Transparency Statement

- All analysis code is publicly available at the GitHub repository: https://github.com/kf120/ibd-biom-glycoda. The data that support the findings of this study are available on reasonable request from the corresponding author.

## Author names in bold designate shared co-first authorship

## Abbreviations

ADCC: antibody-dependent cellular cytotoxicity
AUROC: area under the receiver operating characteristic curve
BAA: biological age acceleration
CDC: complement-dependent cytotoxicity
CD: Crohn’s disease
CLR: centered log-ratio
CMDS: classical multidimensional scaling
CoDA: compositional data analysis
CRP: c-reactive protein
CV: cross-validation
Fc: crystallizable fragment
FCP: fecal calprotectin
FDR: false discovery rate
GP: glycan peak
GlyCmp: GlyCompare-derived glycomotif feature representation
GWAS: genome-wide association studies
HC: healthy controls
IBD: inflammatory bowel disease
IBS: irritable bowel syndrome
IT: Italian cohort
LOCO: leave-one-cohort-out
LR: logistic regression
ML: machine learning
NL: Netherlands cohort
OLS: ordinary least squares
PCA: principal component analysis
SC: symptomatic controls
UC: ulcerative colitis
UHPLC: ultra-high-performance liquid chromatography
UK: United Kingdom cohort
US: United States cohort
XB: XGBoost

---

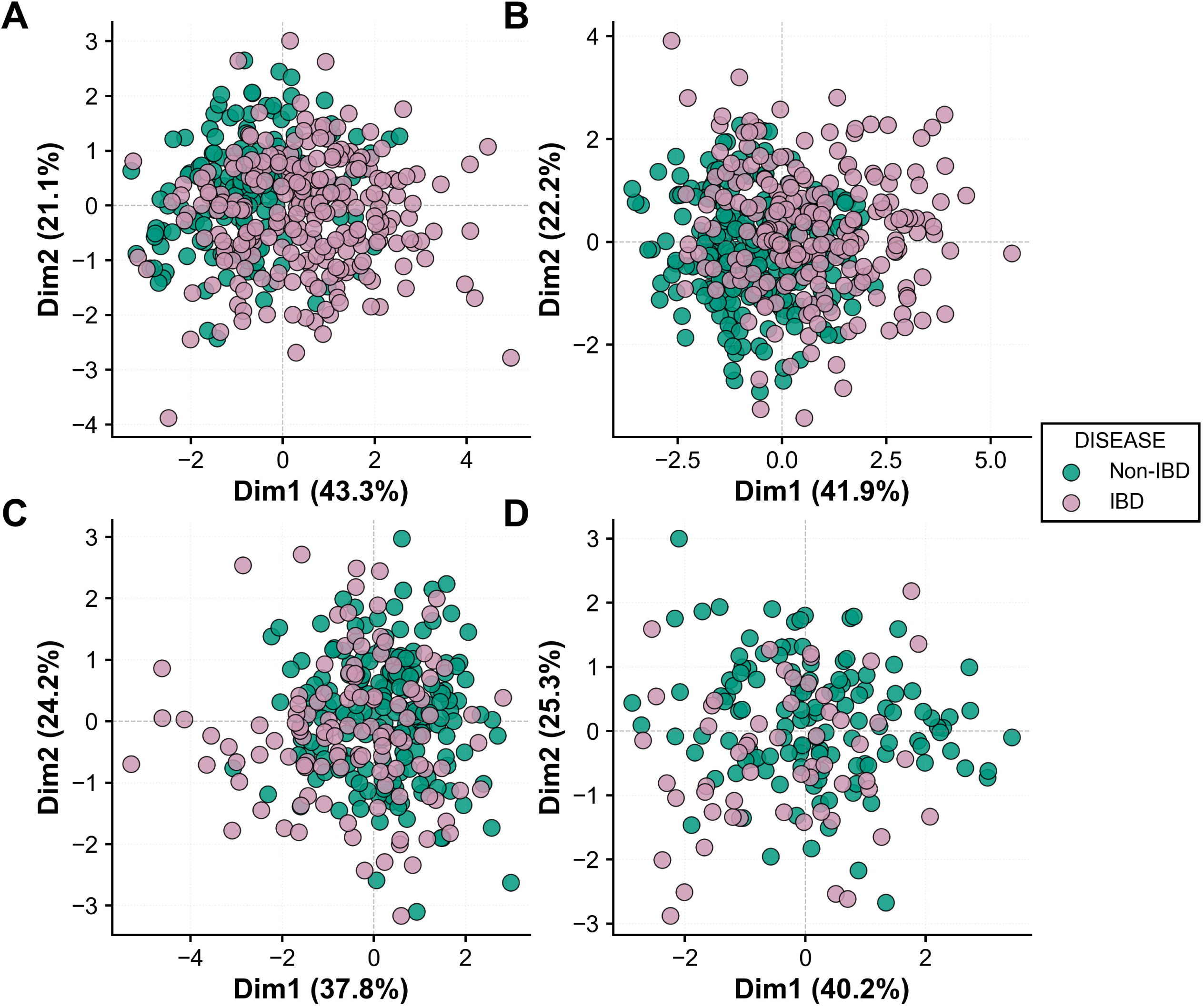

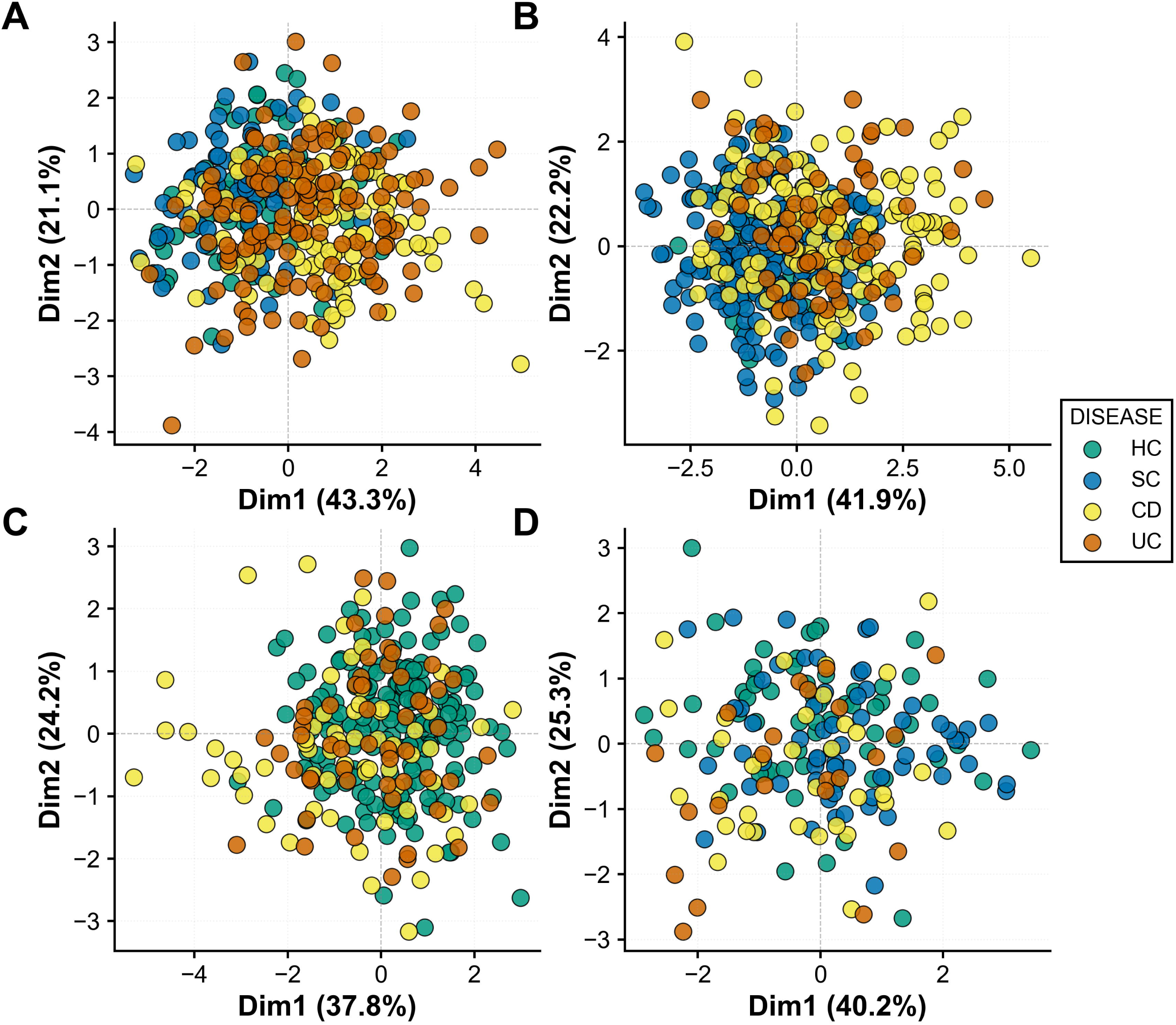

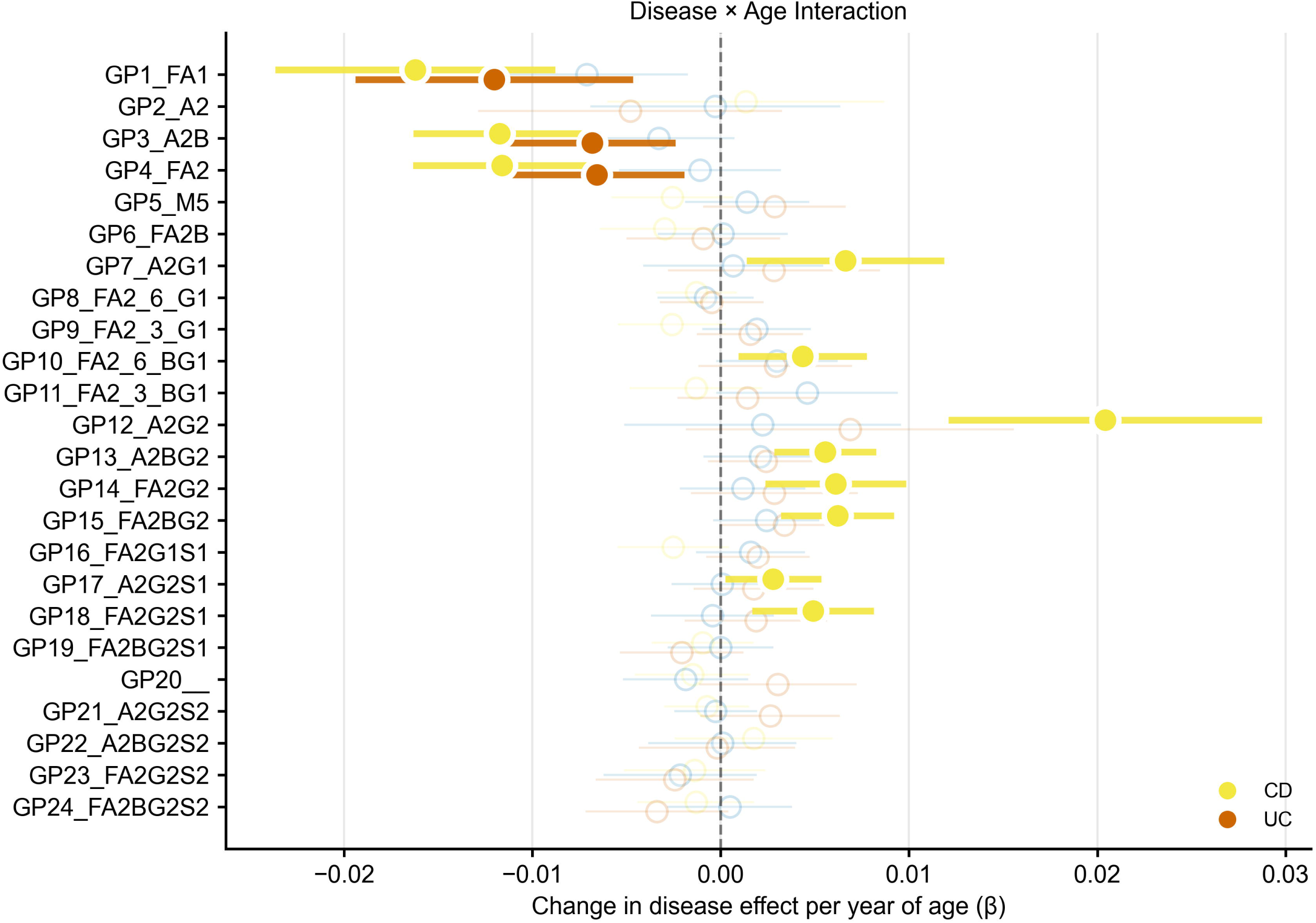

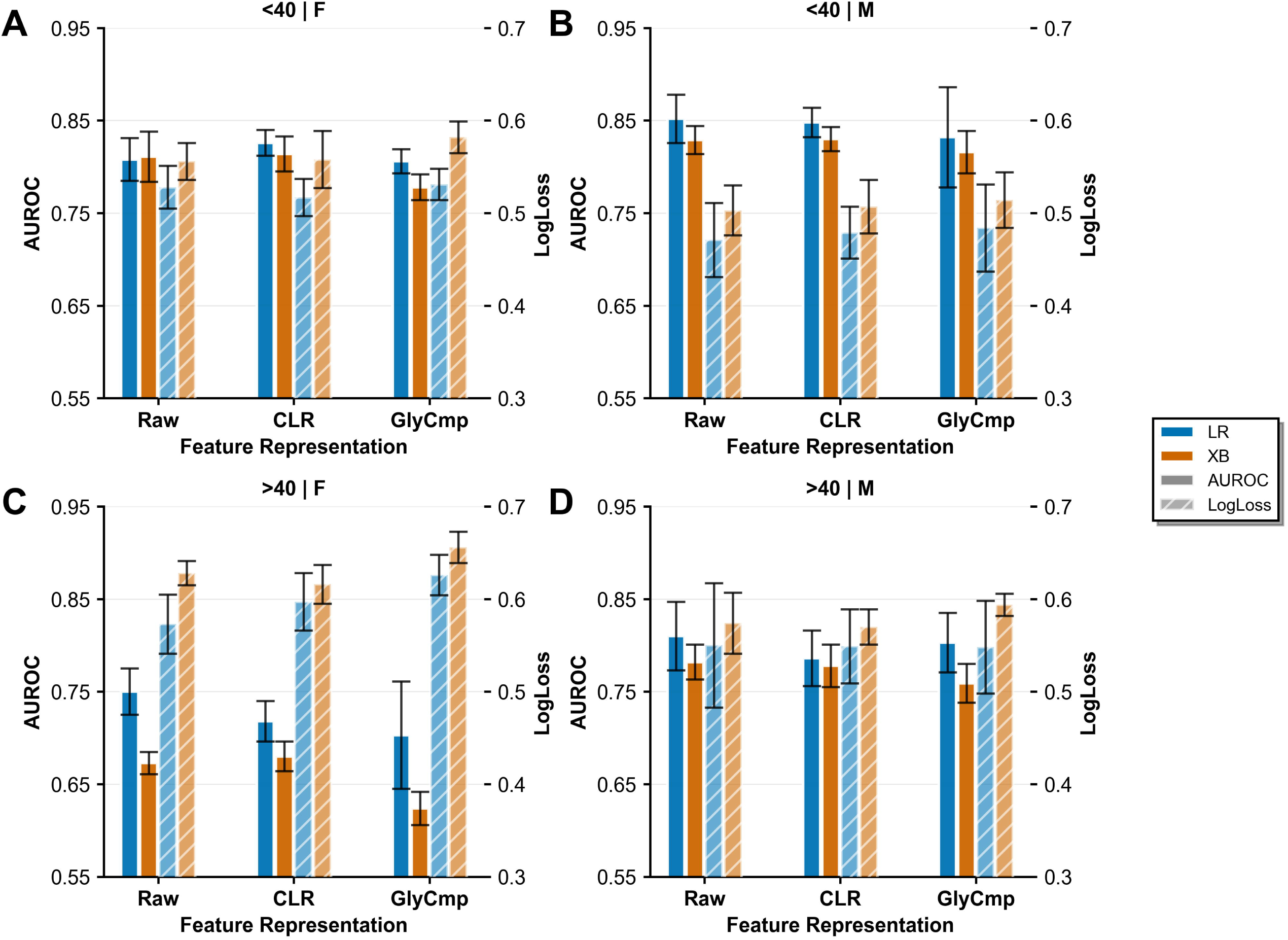

## Notes

### Author Declarations

Tayside Committee on Medical Ethics B gave ethical approval for this work (LREC 06/S1101/16, LREC 2000/4/192); all patients and controls provided written informed consent.

## References

1. Hracs L, Windsor JW, Gorospe J, et al. Global evolution of inflammatory bowel disease across epidemiologic stages. Nature 2025;642:458–466.

2. Kumar A, Yassin N, Marley A, et al. Crossing barriers: the burden of inflammatory bowel disease across Western Europe. Therap Adv Gastroenterol 2023;16:17562848231218615.

3. Spiceland CM, Lodhia N. Endoscopy in inflammatory bowel disease: Role in diagnosis, management, and treatment. World J Gastroenterol 2018;24:4014–4020.

4. Liu D, Saikam V, Skrada KA, et al. Inflammatory Bowel Disease Biomarkers. Med Res Rev 2022;42:1856–1887.

5. Bjarnason I. The Use of Fecal Calprotectin in Inflammatory Bowel Disease. Gastroenterol Hepatol (N Y) 2017;13:53–56.

6. Mumolo MG, Bertani L, Ceccarelli L, et al. From bench to bedside: Fecal calprotectin in inflammatory bowel diseases clinical setting. World J Gastroenterol 2018;24:3681–3694.

7. Chew TS, Mansfield JC. Can faecal calprotectin predict relapse in inflammatory bowel disease: a mini review. Frontline Gastroenterol 2018;9:23–28.

8. Alghoul Z, Yang C, Merlin D. The Current Status of Molecular Biomarkers for Inflammatory Bowel Disease. Biomedicines 2022;10:1492.

9. Mestrovic A, Perkovic N, Bozic D, et al. Precision Medicine in Inflammatory Bowel Disease: A Spotlight on Emerging Molecular Biomarkers. Biomedicines 2024;12:1520.

10. Maverakis E, Kim K, Shimoda M, et al. Glycans In The Immune system and The Altered Glycan Theory of Autoimmunity: A Critical Review. Journal of Autoimmunity 2015;February:1–13.

11. He K, Baniasad M, Kwon H, et al. Decoding the glycoproteome: a new frontier for biomarker discovery in cancer. Journal of Hematology and Oncology 2024;17:12.

12. Rudd PM, Elliott T, Cresswell P, et al. Glycosylation and the immune system. Science 2001;291:2370–2376.

13. Walt D, Aoki-Kinoshita KF, Bendiak B, et al. Transforming Glycoscience: A Roadmap for the Future. Washington, DC: The National Academies Press; 2012.

14. Colley K, Varki A, Haltiwanger R, et al. Cellular Organization of Glycosylation. In: Varki A, Cummings RD, Esko JD, et al., eds. Essentials of Glycobiology. New York: Cold Spring Harbor Laboratory Press; 2022. Available at: https://www.ncbi.nlm.nih.gov/books/NBK579926/.

15. Ohtsubo K, Marth JD. Glycosylation in Cellular Mechanisms of Health and Disease. Cell 2006;126:855–867.

16. Pongracz T, Mayboroda OA, Wuhrer M. The Human Blood N-Glycome: Unraveling Disease Glycosylation Patterns. JACS Au 2024;4:1696–1708.

17. Vidarsson G, Dekkers G, Rispens T. IgG subclasses and allotypes: From structure to effector functions. Frontiers in Immunology 2014;5:1–17.

18. Flevaris K, Kontoravdi C. Immunoglobulin G N-glycan Biomarkers for Autoimmune DiseasesL: Current State and a Glycoinformatics Perspective. International Journal of Molecular Sciences 2022;23:5180.

19. Krištić J, Lauc G. The importance of IgG glycosylation—What did we learn after analyzing over 100,000 individuals. Immunological Reviews 2024.

20. Timoshchuk A, Sharapov S, Aulchenko YS. Twelve Years of Genome-Wide Association Studies of Human Protein N-Glycosylation. Engineering 2023;26:17–31.

21. Dubé R, Rook GAW, Steele J, et al. Agalactosyl IgG in inflammatory bowel disease: Correlation with C-reactive protein. Gut 1990;31:431–434.

22. Shinzaki S, Iijima H, Nakagawa T, et al. IgG oligosaccharide alterations are a novel diagnostic marker for disease activity and the clinical course of inflammatory bowel disease. American Journal of Gastroenterology 2008;103:1173–1181.

23. Trbojevic Akmacic I, Ventham NT, Theodoratou E, et al. Inflammatory bowel disease associates with proinflammatory potential of the immunoglobulin G glycome. Inflammatory Bowel Diseases 2015;21:1237–1247.

24. Simurina M, De Haan N, Vuckovic F, et al. Glycosylation of Immunoglobulin G Associates With Clinical Features of Inflammatory Bowel Diseases. Gastroenterology 2018;154:1320–1333.

25. Gaifem J, Rodrigues CS, Petralia F, et al. A unique serum IgG glycosylation signature predicts development of Crohn’s disease and is associated with pathogenic antibodies to mannose glycan. Nature Immunology 2024. Available at: https://www.nature.com/articles/s41590-024-01916-8.

26. Trbojević-Akmačić I, Lageveen-Kammeijer GSM, Heijs B, et al. High-Throughput Glycomic Methods. Chemical Reviews 2022;122:15865–15913.

27. Haan N de, Pučić-Baković M, Novokmet M, et al. Developments and perspectives in high-throughput protein glycomics: enabling the analysis of thousands of samples. Glycobiology 2022;00:1–13.

28. Bao B, Kellman BP, Chiang AWT, et al. Correcting for sparsity and interdependence in glycomics by accounting for glycan biosynthesis. Nature Communications 2021;12:1–14.

29. Bennett AR, Lundstrøm J, Chatterjee S, et al. Compositional data analysis enables statistical rigor in comparative glycomics. Nature Communications 2025;16:795.

30. Greenacre M, Grunsky E, Bacon-Shone J, et al. Aitchison’s Compositional Data Analysis 40 Years on: A Reappraisal. Statistical Science 2023;38:386–410.

31. Wang J, Lan C, Liu C, et al. Generalizing to Unseen Domains: A Survey on Domain Generalization. IEEE Transactions on Knowledge and Data Engineering 2023;35:8052–8072.

32. Krištić J, Vučković F, Menni C, et al. Glycans are a novel biomarker of chronological and biological ages. Journals of Gerontology - Series A Biological Sciences and Medical Sciences 2014;69:779–789.

33. Ventham NT, Kennedy NA, Nimmo ER, et al. Beyond gene discovery in inflammatory bowel disease: the emerging role of epigenetics. Gastroenterology 2013;145:293–308.

34. Menni C, Keser T, Mangino M, et al. Glycosylation of immunoglobulin G: Role of genetic and epigenetic influences. PLoS ONE 2013;8:6–13.

35. Theodoratou E, Campbell H, Ventham NT, et al. The role of glycosylation in IBD. Nat Rev Gastroenterol Hepatol 2014;11:588–600.

36. Kalla R, Ventham NT, Kennedy NA, et al. MicroRNAs: new players in IBD. Gut 2015;64:504–513.

37. Satsangi J, Kitten O, Chavez M, et al. How to Apply for and Secure EU Funding for Collaborative IBD Research Projects. J Crohns Colitis 2016;10:363–370.

38. Cleynen I, Boucher G, Jostins L, et al. Inherited determinants of Crohn’s disease and ulcerative colitis phenotypes: a genetic association study. Lancet 2016;387:156–167.

39. Vojta A, Dobrinić P, Tadić V, et al. Repurposing the CRISPR-Cas9 system for targeted DNA methylation. Nucleic Acids Res 2016;44:5615–5628.

40. Ventham NT, Kennedy NA, Adams AT, et al. Integrative epigenome-wide analysis demonstrates that DNA methylation may mediate genetic risk in inflammatory bowel disease. Nat Commun 2016;7:13507.

41. Harvey DJ, Merry AH, Royle L, et al. Proposal for a standard system for drawing structural diagrams of N- and O-linked carbohydrates and related compounds. Proteomics 2009;9:3796–3801.

42. Fujita A, Aoki NP, Shinmachi D, et al. The international glycan repository GlyTouCan version 3.0. Nucleic Acids Research 2021;49:D1529–D1533.

43. Thomès L, Burkholz R, Bojar D. Glycowork: A Python package for glycan data science and machine learning. Glycobiology 2021;31:1240–1244.

44. García CB, Salmerón R, García C, et al. Residualization: justification, properties and application. J Appl Stat 47:1990–2010.

45. Wang J. Classical Multidimensional Scaling. In: Wang J, ed. Geometric Structure of High-Dimensional Data and Dimensionality Reduction. Berlin, Heidelberg: Springer; 2012:115–129. Available at: 10.1007/978-3-642-27497-8_6.

46. Seabold S, Perktold J. Statsmodels: Econometric and Statistical Modeling with Python. In: Austin, Texas; 2010:92–96. Available at: https://doi.curvenote.com/10.25080/Majora-92bf1922-011.

47. Virtanen P, Gommers R, Oliphant TE, et al. SciPy 1.0: fundamental algorithms for scientific computing in Python. Nature Methods 2020;17:261–272.

48. Zhang Y, Krishnan S, Bao B, et al. Preparing glycomics data for robust statistical analysis with GlyCompareCT. STAR Protocols 2023;4:102162.

49. Chen T, Guestrin C. XGBoost: A Scalable Tree Boosting System. In: KDD ’16: Proceedings of the 22nd ACM SIGKDD International Conference on Knowledge Discovery and Data Mining.

50. Filho TS, Song H, Perello-Nieto M, et al. Classifier Calibration: How to assess and improve predicted class probabilities: a survey. Machine Learning 2023. Available at: http://arxiv.org/abs/2112.10327.

51. Pedregosa F, Varoquaux G, Gramfort A, et al. Scikit-learn: Machine Learning in Python. Journal of Machine Learning Research 2011;12:2825–2830.

52. Shkunnikova S, Mijakovac A, Sironic L, et al. IgG glycans in health and disease: Prediction, intervention, prognosis, and therapy. Biotechnology advances 2023:108169.

53. Rapčan B, Song M, Frkatović-Hodžić A, et al. Glycan clock of ageing-analytical precision and time-dependent inter- and i-individual variability. Geroscience 2024;46:5781–5796.

54. Vinicki M, Pribić T, Vučković F, et al. Effects of testosterone and metformin on the GlycanAge index of biological age and the composition of the IgG glycome. GeroScience 2024;47:1777–1788.

55. Levine ME, Lu AT, Quach A, et al. An epigenetic biomarker of aging for lifespan and healthspan. Aging (Albany NY) 2018;10:573–591.

56. Cao B, Zhao X, Lu Z, et al. Accelerated biological aging and risk of inflammatory bowel disease: A prospective study from 401,013 participants. J Nutr Health Aging 2025;29:100505.

57. Lu AT, Quach A, Wilson JG, et al. DNA methylation GrimAge strongly predicts lifespan and healthspan. Aging (Albany NY) 2019;11:303–327.

58. Ventham NT, Kennedy NA, Kalla R, et al. Genome-Wide Methylation Profiling in 229 Patients With Crohn’s Disease Requiring Intestinal Resection: Epigenetic Analysis of the Trial of Prevention of Post-operative Crohn’s Disease (TOPPIC). Cell Mol Gastroenterol Hepatol 2023;16:431–450.

59. Noble A, Adams A, Nowak J, et al. The Circulating Methylome in Childhood-Onset Inflammatory Bowel Disease. J Crohns Colitis 2025;19:jjae157.

60. Ramsteijn A, Noble A, Antunes C, et al. P1309 Biological age acceleration in Crohn’s Disease patients in long-term remission on combination therapy: epigenetic analysis of the SPARE & STORI trial population. J Crohns Colitis 2026;20:jjaf231.1490.

61. Vučkovïc F, Krištïc J, Gudelj I, et al. Association of systemic lupus erythematosus with decreased immunosuppressive potential of the IgG glycome. Arthritis and Rheumatology 2015;67:2978–2989.

62. Lemmers RFH, Vilaj M, Urda D, et al. IgG glycan patterns are associated with type 2 diabetes in independent European populations. Biochimica et Biophysica Acta - General Subjects 2017;1861:2240–2249.

63. Desai K, Gupta S, May FP, et al. Early Detection of Advanced Adenomas and Colorectal Carcinoma by Serum Glycoproteome Profiling. Gastroenterology 2024;166:194–197.e2.

64. Flevaris K, Davies J, Nakai S, et al. Machine learning framework to extract the biomarker potential of plasma IgG N-glycans towards disease risk stratification. Computational and Structural Biotechnology Journal 2024;23:1234–1243.

65. Iacucci M, Santacroce G, Yasuharu M, et al. Artificial Intelligence–Driven Personalized Medicine: Transforming Clinical Practice in Inflammatory Bowel Disease. Gastroenterology 2025;169:416–431.

66. El-Sayed A, Lovat LB, Ahmad OF. Clinical Implementation of Artificial Intelligence in Gastroenterology: Current Landscape, Regulatory Challenges, and Ethical Issues. Gastroenterology 2025;169:518–530.

67. Trbojević-Akmačić I, Vučković F, Pribić T, et al. Comparative analysis of transferrin and IgG N-glycosylation in two human populations. Commun Biol 2023;6:312.

68. Clerc F, Novokmet M, Dotz V, et al. Plasma N-Glycan Signatures Are Associated With Features of Inflammatory Bowel Diseases. Gastroenterology 2018;155:829–843.

