## Supplemental Material for "A Multi-Cohort Study of Immunoglobulin G Glycans in Newly Diagnosed Inflammatory Bowel Disease Patients Reveals Accelerated Biological Aging"

### **Title**

### **Short Title**

IgG N-glycans Reveal Accelerated Biological Aging in IBD

### **Author(s)**

Konstantinos Flevaris^1^*, Irena Trbojević-Akmačić^2^*, David Goh^1^, Juproop Singh Lalli^1^, Frano Vučković^2^, Marija Ćapin Vilaj^2^, Jerko Štambuk^2^, Jasminka Krištić^2^, Anika Mijakovac^2,3^, Nick Ventham^4^, Rahul Kalla^5^, Anna Latiano^6^, Natalia Manetti^7^, Dalin Li^8^, Dermot P.B. McGovern^8^, Nicholas A. Kennedy^9^, Vito Annese^10^, Gordan Lauc^2,11^**, Jack Satsangi^12^**, Cleo Kontoravdi^1^**

* shared first authors; ** shared last authors

**Affiliations:**

^1^Department of Chemical Engineering, Imperial College London, London, UK

^2^Genos Glycoscience Research Laboratory, Zagreb, Croatia

^3^University of Zagreb, Faculty of Science, Zagreb, Croatia

^4^Edinburgh IBD Unit, Western General Hospital, Edinburgh, UK

^5^ Centre for Inflammation Research, Institute of Regeneration and Repair, University of Edinburgh, Edinburgh, UK

^6^Division of Gastroenterology, Fondazione IRCCS - Casa Sollievo della Sofferenza, San Giovanni Rotondo, Italy

^7^Gastroenterology Unit, San Jacopo Hospital (Pistoia), USL Toscana Centro

^8^F. Widjaja Inflammatory Bowel Disease Institute, Department of Medicine, Cedars-Sinai Medical Centre, Los Angeles, CA, USA

^9^Exeter Biomedical Research Centre, University of Exeter, UK

^10^Vita salute University San Raffaele, Milan - Head of Gastroenterology IRCCS Policlinic San Donato Milanese, MI, Italy

^11^University of Zagreb, Faculty of Pharmacy and Biochemistry, Zagreb, Croatia

^12^Translational Gastroenterology & Hepatology Unit, Nuffield Department of Medicine University of Oxford, UK

**Calculation of GlycanAge index**

Biological age was quantified based on the GlycanAge index, which was calculated using the following linear model in *R*:

*lm(Age ~ poly(GP6, 2, raw = TRUE) × Sex + GP14 × Sex + GP15 × Sex)*

Model coefficients were trained on a reference set of 335 HC. GlycanAge values were subsequently predicted using this model for both controls and cases (i.e., CD and UC).

### Table S1. Summary of IgG N-glycan peaks (GPs) analyzed in this study

| **GP** | **Oxford Notation** | **GlyTouCan ID** | **Description** |
| --- | --- | --- | --- |
| GP1 | FA1 | G88876JQ | Monoantennary, agalactosylated, core fucosylated |
| GP2 | A2 | G88876JQ | Biantennary, agalactosylated, non-fucosylated |
| GP3 | A2B | G76241TN / G44754DF | Biantennary, agalactosylated, bisected, non-fucosylated |
| GP4 | FA2 | G65984FE | Biantennary, agalactosylated, core fucosylated |
| GP5 | M5 | G60965VN | Biantennary, high-mannose, non-fucosylated |
| GP6 | FA2B | G60965VN / G53582JE | Biantennary, agalactosylated, bisected, core fucosylated |
| GP7 | A2G1 | G36859SD / G44754DF |  |
| GP8 | FA2[6]G1 | G44754DF | Biantennary, monogalactosylated (a6-antenna), core fucosylated |
| GP9 | FA2[3]G1 | G36859SD | Biantennary, monogalactosylated (a3-antenna), core fucosylated |
| GP10 | FA2[6]BG1 | G92575UZ | Biantennary, monogalactosylated (a6-antenna), bisected, core fucosylated |
| GP11 | FA2[3]BG1 | G20079RG | Biantennary, monogalactosylated (a3-antenna), bisected, core fucosylated |
| GP12 | A2G2 | G66741YQ |  |
| GP13 | A2BG2 | G36522OD | Biantennary, digalactosylated, bisected, non-fucosylated |
| GP14 | FA2G2 | G00998NI |  |
| GP15 | FA2BG2 | G74325TG | Biantennary, digalactosylated, bisected, core fucosylated |
| GP16 | FA2G1S1 | G71782CJ / G19447ZX | Biantennary, monogalactosylated, monosialylated, core fucosylated |
| GP17 | A2G2S1 | G01670UQ / G37591JC | Biantennary, digalactosylated, monosialylated, non-fucosylated |
| GP18 | FA2G2S1 | G01361CV / G83424BL | Biantennary, digalactosylated, monosialylated, core fucosylated |
| GP19 | FA2BG2S1 | G42295MA | Biantennary, digalactosylated, bisected, monosialylated, core fucosylated |
| GP20 | - | - | Structure not determined |
| GP21 | A2G2S2 | - | Biantennary, digalactosylated, disialylated, non-fucosylated |
| GP22 | A2BG2S2 | G14854WL | Biantennary, digalactosylated, bisected, disialylated, non-fucosylated |
| GP23 | FA2G2S2 | G72954XC | Biantennary, digalactosylated, disialylated, core fucosylated |
| GP24 | FA2BG2S2 | G48713OY | Biantennary, digalactosylated, bisected, disialylated, core fucosylated |

#### Table S2. Clustering metrics (Dunn index and mean silhouette width) by disease status labeling (binary vs. multiclass) in UK, US, IT, and NL cohorts

| **Cohort** | **Binary –**  **Dunn Index** | **Binary –**  **Silhouette Width** | **Multiclass - Dunn Index** | **Multiclass - Silhouette Width** |
| --- | --- | --- | --- | --- |
| UK | 0.078 | 0.064 | 0.078 | -0.033 |
| US | 0.074 | 0.078 | 0.074 | -0.042 |
| IT | 0.099 | 0.047 | 0.099 | 0.019 |
| NL | 0.128 | 0.046 | 0.126 | -0.023 |

#### Table S3. Hyperparameters used for logistic regression (LR) and XGBoost (XB) in this study

| **Hyperparameter** | **Logistic Regression (LR)** | **XGBoost (XB)** |
| --- | --- | --- |
| C | 1 | — |
| penalty | l2 | — |
| solver | lbfgs | — |
| n_estimators | — | 100 |
| learning_rate | — | 0.01 |
| max_depth | — | 2 |
| min_child_weight | — | 10 |
| early_stopping_rounds | — | 5 |

#### Table S4. Overall absolute out-of-sample performance for binary classification of IBD (CD and UC) vs. Non-IBD (HC and SC) across LOCO-CV runs for ML pipelines. Reported metric values represent pooled means across runs, with 95% confidence intervals shown in parentheses.

| **Pipeline** | **AUROC** | **Log Loss** | **Sensitivity** | **Specificity** | **Worst Cohort** |
| --- | --- | --- | --- | --- | --- |
| LR+Raw | 0.799 (0.726, 0.872) | 0.536 (0.449, 0.622) | 0.651 (0.588, 0.714) | 0.792 (0.679, 0.905) | IT |
| LR+CLR | 0.791 (0.735, 0.848) | 0.538 (0.449, 0.626) | 0.680 (0.583, 0.776) | 0.752 (0.654, 0.850) | IT |
| LR+GlyCmp | 0.778 (0.713, 0.842) | 0.554 (0.495, 0.612) | 0.636 (0.578, 0.695) | 0.779 (0.684, 0.874) | IT |
| XB+Raw | 0.762 (0.677, 0.847) | 0.568 (0.527, 0.610) | 0.613 (0.535, 0.690) | 0.786 (0.710, 0.863) | IT |
| XB+CLR | 0.763 (0.703, 0.823) | 0.564 (0.520, 0.609) | 0.612 (0.543, 0.680) | 0.799 (0.733, 0.865) | IT |
| XB+GlyCmp | 0.740 (0.697, 0.782) | 0.586 (0.561, 0.612) | 0.593 (0.513, 0.673) | 0.762 (0.691, 0.832) | IT |

#### Table S5. Absolute out-of-sample performance for binary classification of IBD (CD and UC) vs. Non-IBD (HC and SC) across LOCO-CV runs, focusing on females younger than 40 years. Reported metric values represent pooled averages across runs, with 95% confidence intervals shown in parentheses.

| **Pipeline** | **AUROC** | **Log Loss** |
| --- | --- | --- |
| LR+Raw | 0.808 (0.734, 0.882) | 0.528 (0.455, 0.601) |
| LR+CLR | 0.826 (0.780, 0.872) | 0.517 (0.452, 0.582) |
| LR+GlyCmp | 0.806 (0.763, 0.849) | 0.531 (0.475, 0.586) |
| XB+Raw | 0.811 (0.727, 0.896) | 0.556 (0.491, 0.621) |
| XB+CLR | 0.814 (0.755, 0.873) | 0.558 (0.459, 0.657) |
| XB+GlyCmp | 0.778 (0.735, 0.821) | 0.582 (0.526, 0.638) |

#### Table S6. Absolute out-of-sample performance for binary classification of IBD (CD and UC) vs. Non-IBD (HC and SC) across LOCO-CV runs, focusing on males younger than 40 years. Reported metric values represent pooled averages across runs, with 95% confidence intervals shown in parentheses.

| **Pipeline** | **AUROC** | **Log Loss** |
| --- | --- | --- |
| LR+Raw | 0.852 (0.770, 0.934) | 0.471 (0.344, 0.599) |
| LR+CLR | 0.848 (0.798, 0.897) | 0.479 (0.390, 0.568) |
| LR+GlyCmp | 0.832 (0.661, 1.003) | 0.484 (0.335, 0.634) |
| XB+Raw | 0.829 (0.780, 0.878) | 0.503 (0.417, 0.588) |
| XB+CLR | 0.830 (0.789, 0.871) | 0.507 (0.413, 0.600) |
| XB+GlyCmp | 0.816 (0.743, 0.889) | 0.514 (0.417, 0.611) |

#### Table S7. Absolute out-of-sample performance for binary classification of IBD (CD and UC) vs. Non-IBD (HC and SC) across LOCO-CV runs, focusing on females older than 40 years. Reported metric values represent pooled averages across runs, with 95% confidence intervals shown in parentheses.

| **Pipeline** | **AUROC** | **Log Loss** |
| --- | --- | --- |
| LR+Raw | 0.750 (0.671, 0.830) | 0.573 (0.470, 0.676) |
| LR+CLR | 0.718 (0.648, 0.788) | 0.597 (0.497, 0.696) |
| LR+GlyCmp | 0.703 (0.518, 0.888) | 0.626 (0.555, 0.698) |
| XB+Raw | 0.673 (0.634, 0.712) | 0.628 (0.585, 0.671) |
| XB+CLR | 0.680 (0.628, 0.732) | 0.616 (0.549, 0.682) |
| XB+GlyCmp | 0.624 (0.568, 0.680) | 0.656 (0.602, 0.710) |

#### Table S8. Absolute out-of-sample performance for binary classification of IBD (CD and UC) vs. Non-IBD (HC and SC) across LOCO-CV runs, focusing on males older than 40 years. Reported metric values represent pooled averages across runs, with 95% confidence intervals shown in parentheses.

| **Pipeline** | **AUROC** | **Log Loss** |
| --- | --- | --- |
| LR+Raw | 0.810 (0.694, 0.927) | 0.550 (0.336, 0.764) |
| LR+CLR | 0.786 (0.690, 0.881) | 0.549 (0.423, 0.675) |
| LR+GlyCmp | 0.803 (0.700, 0.906) | 0.548 (0.390, 0.706) |
| XB+Raw | 0.782 (0.722, 0.843) | 0.574 (0.469, 0.679) |
| XB+CLR | 0.778 (0.705, 0.851) | 0.570 (0.511, 0.629) |
| XB+GlyCmp | 0.759 (0.693, 0.824) | 0.594 (0.554, 0.633) |

#### Table S9. Overall absolute out-of-sample performance for binary classification of CD vs. UC across LOCO-CV runs for LR pipelines. Reported metric values represent pooled means across runs, with 95% confidence intervals shown in parentheses.

| **Pipeline** | **AUROC** | **Log Loss** | **Sensitivity** | **Specificity** | **Worst Cohort** |
| --- | --- | --- | --- | --- | --- |
| LR+Raw | 0.726 (0.581, 0.870) | 0.617 (0.494, 0.740) | 0.622 (0.397, 0.848) | 0.729 (0.65, 0.808) | NL |
| LR+CLR | 0.723 (0.591, 0.854) | 0.618 (0.498, 0.739) | 0.659 (0.481, 0.838) | 0.700 (0.574, 0.826) | NL |
| LR+GlyCmp | 0.701 (0.383, 1.018) | 0.664 (0.655, 0.673) | 0.581 (0.387, 0.776) | 0.741 (0.666, 0.815) | NL |

#### Figure S1. Similarity analysis of IgG N-glycan profiles across disease groups and cohorts using biplots of CLR-transformed and residualized GP profiles for Non-IBD and IBD individuals across (A) UK, (B) US, (C) IT, and (D) NL cohorts. Each point represents an individual clinical sample. The first two principal components (PC1 and PC2) are shown on the x- and y-axes, respectively, with the percentage of variance explained indicated in parentheses.

#### Figure S2. Similarity analysis of IgG N-glycan profiles across disease groups and cohorts using biplots of CLR-transformed and residualized GP profiles for HC, SC, CD, and UC individuals across (A) UK, (B) US, (C) IT, and (D) NL cohorts. Each point represents an individual clinical sample. The first two principal components (PC1 and PC2) are shown on the x- and y-axes, respectively, with the percentage of variance explained indicated in parentheses.

### Figure S3. Age-related modification of disease-associated differences in IgG N-glycans in SC and IBD. Lollipop plot showing the coefficients for the disease × age interaction from linear models for each CLR-transformed GP. Estimates are shown for SC, CD, and UC relative to HC. Points indicate interaction-effect estimates and horizontal lines denote 95% confidence intervals. Significant interactions (FDR-adjusted p < 0.05) are shown prominently, whereas non-significant interactions are faded. Positive values indicate that the disease effect becomes stronger with increasing age, whereas negative values indicate attenuation with age.

### Figure S4. Subgroup-specific out-of-sample performance for ML prediction models classifying Non-IBD vs. IBD under LOCO-CV. Absolute out-of-sample performance in terms of AUROC and log loss is shown across all held-out cohorts for each feature representation method (raw GPs, CLR-transformed GPs, or GlyCmp-transformed GPs) and model class (LR or XB) for (A) females aged ≤40 years, (B) males aged ≤40 years, (C) females aged >40 years, and (D) males aged >40 years. Each bar represents the mean out-of-sample performance across held-out cohorts for a given model class and feature representation. Colors indicate model class, hatching indicates the evaluation metric, and error bars denote the standard error across held-out cohorts.
